# Long-term spatial patterns in COVID-19 booster vaccine uptake

**DOI:** 10.1101/2022.08.30.22279415

**Authors:** A.J. Wood, A.M. MacKintosh, M. Stead, R.R. Kao

## Abstract

Vaccination is a critical tool for controlling infectious diseases, with its use to protect against COVID-19 being a prime example. Where a disease is highly transmissible, even a small proportion of unprotected individuals can have substantial implications for disease burdens and compromise disease control. As socio-demographic factors such as deprivation and ethnicity have been shown to influence uptake rates, identifying how uptake varies with socio-demographic indicators is a critical step for reducing hesitancy and issues of access, and identifying plausible future uptake patterns.

Here, we analyse the numbers of COVID-19 booster vaccinations subdivided by age, sex, dose and location. We use publicly available socio-demographic data, and use Random Forest models to capture patterns of first booster uptake at high spatial resolution, with systematic variation restricted to *∼*1km in urban areas. We introduce a novel method to predict future distributions using our first booster model, assuming existing trends with respect to deprivation will persist. This provides a quantitative estimate of the impact of changing motivations and efforts to increase uptake.

While age and sex have the greatest impact on the model fit, there is a substantial influence of community deprivation and the proportion of residents belonging to a black or minority ethnicity. Changes in patterns from first to second boosters suggest in the longer-term that the impact of deprivation is likely to increase, furthering the disproportionate impact of COVID-19 on deprived communities. Our analysis is based solely on publicly available socio-demographic data and readily recorded vaccination data, and would be easily adaptable to uptake data from countries where data recording is similar, and for aiding vaccination campaigns against other infectious diseases.

## 1 Introduction

Vaccine hesitancy is a critical problem that severely impacts our ability to control important infectious diseases such as measles and seasonal influenza, and has been the subject of much scrutiny during the COVID-19 pandemic. In a voluntary campaign, uptake will depend on individual decision-making and this is known to be influenced by sociological and demographic factors [1, 2]. Quantifying these factors can be challenging but is invaluable for understanding the context of individual decisions and developing strategies to improve rates of uptake.

Scotland’s COVID-19 vaccination programme began in December 2020 and as of May 2024 had delivered over 14 million doses to a population of 5.4 million, with about 93% of those aged 20+ receiving at least one dose. A primary course for all adults was followed by subsequent rounds of boosters, the first in Autumn/Winter 2021 primarily targeted at ages 50+ [3], but rapidly expanded to all adults in response to a wave of the B.1.1.529 *Omicron* variant of concern (VOC) in November 2021 [4]. Regular rounds of booster vaccination have taken place since, but aimed at specific age groups and those otherwise considered vulnerable to severe disease, first in Spring 2022 for all aged 75+ [5], then in Autumn 2022 with all aged 50+ eligible [6], and similar rounds every six months thereafter. The immunity against severe disease provided from a booster typically lasts less than a year [7, 8]. The pressure on healthcare systems in Scotland from COVID-19 has remained substantial; most recently a wave of infection in Summer 2024 (monitored primarily through wastewater surveillance) preceded an increase in COVID-19 related hospital admissions, peaking at 485 in week ending July 7 2024 (contrasting with an all-time peak of 1069 in w/e April 7 2020) [9]. Waning immunity and the narrower eligibility criteria for future boosters raises the question of how strategies aimed at increasing uptake should be targeted, especially with respect to age and deprivation which are both significant risk factors for severe COVID-19 outcomes [10, 11, 12, 13].

Belonging to a black or other minority ethnicity and living in a community with more severe socioeconomic deprivation are significant and consistent risk factors for lower uptake of routine vaccinations (e.g. shingles [14, 15] and seasonal influenza in adults [16, 14, 17], and seasonal influenza [18, 16], HPV [19] and MMR [20] in children when considering parents’ status). Socioeconomic deprivation is usually presented in these studies in terms of an ‘overall’ deprivation quintile of the community an individual resides in, aggregating measures over many different aspects of deprivation (e.g. income, education, health) [21, 22]. At the individual level for COVID-19 vaccination, attitudes towards accepting follow-up booster doses over time may differ from that towards the initial course of vaccination, especially as the perceived threat of COVID-19 changes over time. Surveys on hesitancy have highlighted reasons for *accepting* the first vaccination course that were specific to the context of the pandemic at that time. These include a desire for life to get “back to normal”, a feeling of moral duty, and concern of potential requirements of vaccination to travel [23, 24, 25, 26, 27]. Hesitancy varied over the early stages of the epidemic [28, 29, 30] and in Scotland uptake has fallen on each successive round of vaccination. It is reasonable to expect uptake to fall further in the future, barring radical changes in pathogen virulence or transmissibility. Our aims with this paper are twofold. First, to describe for the first and second booster programmes the variation in uptake across demographics and specific markers for socioeconomic deprivation, and show how that manifests as a spatial clustering of communities with low uptake. For first boosters in particular we use a Random Forest regression model to explain spatial variation in uptake using known risk factors for vaccine hesitancy, refusal and access, thereby quantifying the importance of these risk factors in a geographically explicit context. Our second aim is to explore a novel method for using this model for first boosters to *predict* [31] future spatial variation, should uptake continue to fall and the risk factors for vaccine hesitancy were to persist. Owing to the changing eligibility criteria and the context of the pandemic since the first booster programme, we should not expect these predictions to line up with second boosters and beyond, but it does allow us to quantify the effect of these changing circumstances.

## 2 Data

Scottish vaccination data are provided by Public Health Scotland’s electronic Data Research and Innovation Service (eDRIS), under a data sharing agreement. Individuals are grouped by sex, age range (0–19, 20–29, 30–39, 40–49, 50–59, 60–69, 70+), and residing datazone (DZ; census areas of order 500–1,000 individuals, each with an area as low as 0.15–0.4km^2^ in densely populated areas). For each of these groups, the data give the number of individuals to have received exactly 1 dose, exactly 2 doses, exactly 3 doses, and exactly 4 doses. When the number of individuals is fewer than 5, the exact number is not provided, and we take an estimate (see Supplementary Material, Section A).

Population denominators are taken from census table UV102b, giving small-area populations by age and sex as of 22 March 2022 [32]. Data on small-area population breakdown by ethnicity are also obtained from 2022 census data, table UV201b [33].

Measures of deprivation are taken from the *Scottish Index of Multiple Deprivation* (SIMD) dataset [22]. This contains measures of different indicators of deprivation at DZ level (e.g., the percentage of residents living in overcrowded housing). The SIMD also ranks DZs by deprivation in each of *access*, *income*, *employment*, *education*, *health*, *crime* and *housing*. These ranks are derived from a weighted average of individual deprivation measures (see Supplementary Material, Section A for further details). A DZ with rank 1 is considered to have the highest relative deprivation, and a DZ with rank 6,976 (out of 6,976) the lowest. An overall deprivation rank and decile are also given from a weighted average of all measures.

COVID-19 vaccination in Scotland began with administration of a first primary dose, and a second primary dose from eight weeks after. Three months from this initial course, adults then become eligible for a first booster dose, commencing Autumn 2021. A second round of booster vaccination was available to over 75’s and those otherwise considered vulnerable to severe COVID-19 disease in Spring 2022. Then a further round of booster vaccination was available to over 50’s and those otherwise vulnerable in Autumn 2022.

We distinguish between two characterisations of booster uptake:

- *Overall* uptake is the proportion of individuals to have received a booster vaccination. The denominator is the population.
- *Returning* uptake is the proportion of individuals *that have received at least one dose*, to have returned for a booster. The denominator is the number of individuals to have received at least one dose. The product of returning booster uptake and overall first dose uptake is then the overall booster uptake.

Our model is fit to returning uptake but we report in terms of overall uptake where appropriate.

We exclude the 0–19 age bracket which includes many very young individuals who were not eligible for any vaccine or booster. Finally, a small fraction of individuals with severely weakened immune systems are eligible for additional primary doses, on top of boosters [34]. Due to the structure of the data used here, we define first booster uptake for all individuals as uptake of the third available dose, of any type, and second booster uptake as uptake of the fourth available dose.

## 3 Distributions of first and second booster uptake

First and second booster uptakes are summarised in Fig 1 with respect to age, sex, and deprivation decile. First booster uptake was 79% in ages 20+ across Scotland overall (returning after a first dose: 85%) and 90% in ages 50+ overall (returning: 93%). Uptake was lower in younger, more deprived subpopulations, as well as in men compared to women. The oldest groups sustained high uptake at all deprivation indices.

**Figure 1:**
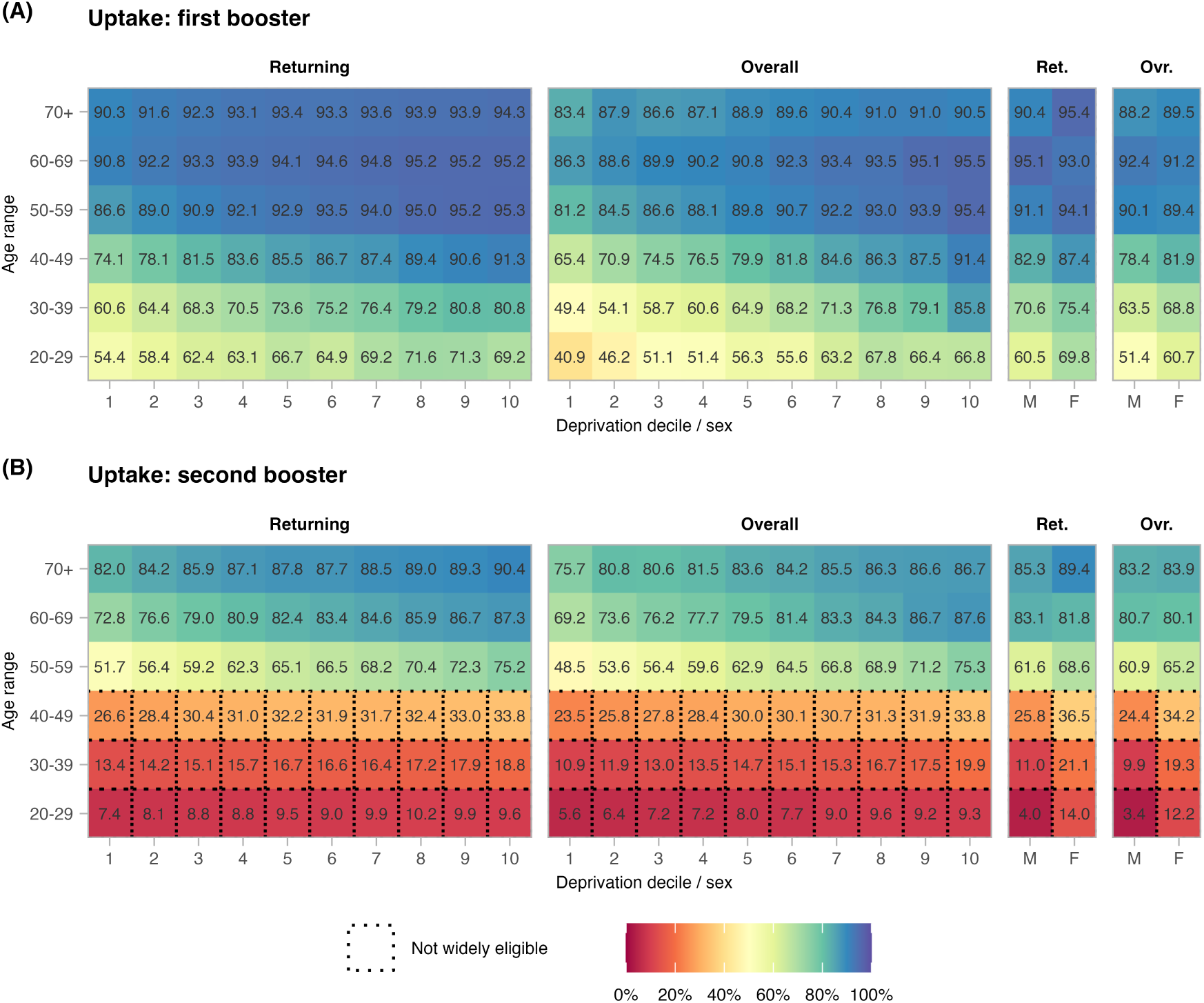
Booster vaccination uptake with respect to age range, sex and DZ deprivation decile, comparing (**A**) first boosters to (**B**) second boosters. “Returning” uptake is the proportion of individuals to have returned for a booster, of those that have received at least one dose previously. “Overall” uptake is the proportion of all individuals to have received a booster (the denominator being the population size). Decile 1 contains the most deprived DZs, and decile 10 the least deprived. Second boosters were not widely available to those aged below 50.

For second boosters, focusing on ages 50+ (ages *<*50 not being widely eligible), uptake fell to 75% (returning: 78%). The skews across each of age, sex and deprivation persist in second booster uptake. We quantify the change in skew with the ratio of returning uptake between *(i)* men and women, *(ii)* deprivation deciles 1 and 10, *(iii)* ages 50–59 and 70+. Comparing first booster to second booster, the skew in sex increased from 1.02 to 1.05, in deprivation increased from 1.07 to 1.26, and in age increased from 1.01 to 1.34. Thus as well as uptake declining from first to second booster, inequalities in each of these metrics were further exacerbated, especially across age and deprivation.

Figure 2A shows deprivation percentiles over the densely populated *central belt* of Scotland, and Figures 2B and C respectively show uptake for first and second boosters. The skews in uptake with respect to deprivation manifest as a spatial clustering of areas with lower uptake, coinciding with communities with higher deprivation.

**Figure 2:**
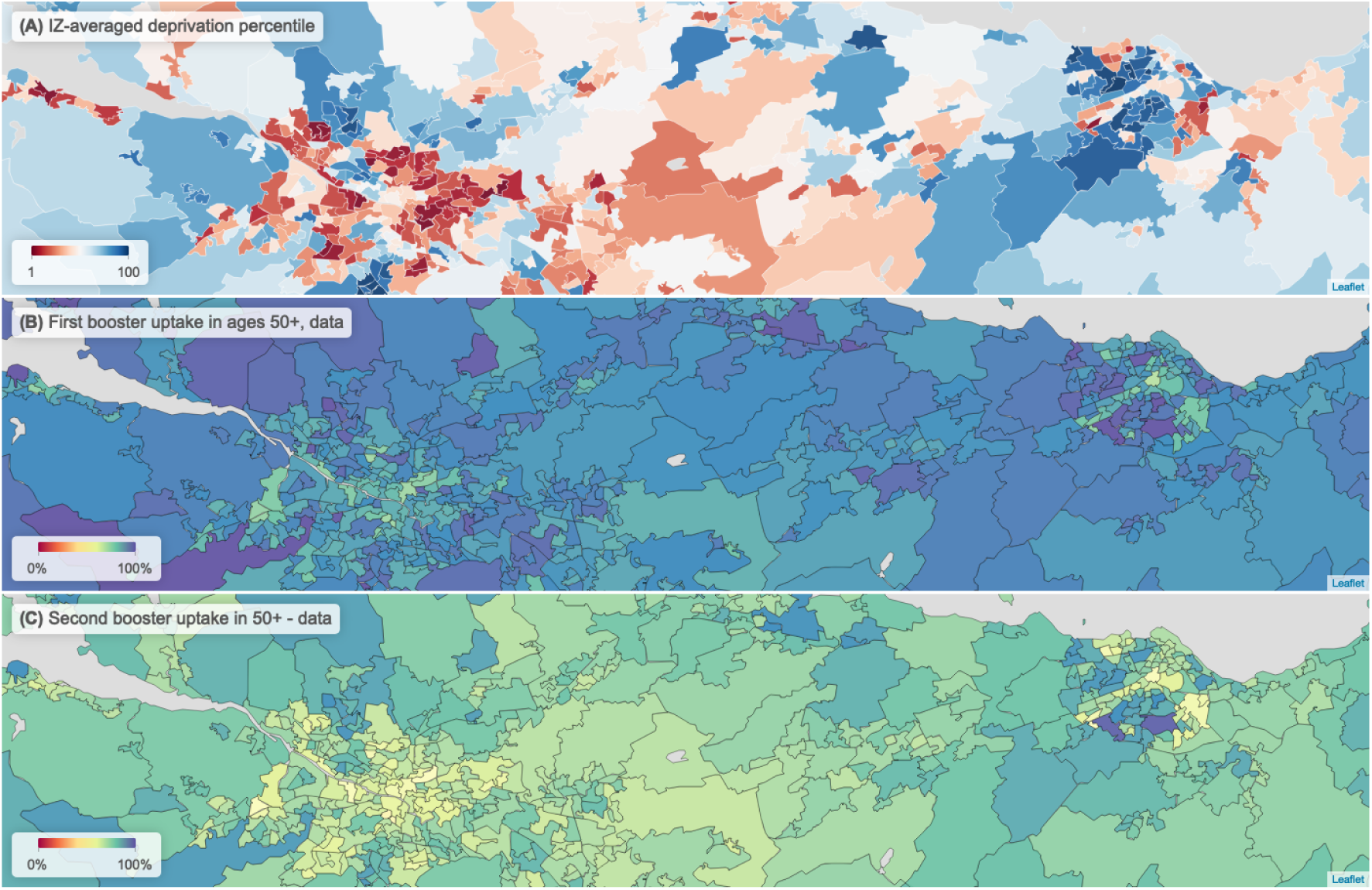
Map view of (**A**) deprivation, (**B**) overall 50+ first booster uptake, and (**C**) overall 50+ second booster uptake across the *central belt* of Scotland, containing the cities of Glasgow (left cluster) and Edinburgh (right cluster). These are presented at *intermediate zone* level; geographical areas typically containing 4–6 DZs. Clusters of higher deprivation coincide with clusters of low uptake across both first and second boosters.

### 3.1 A model for first booster uptake

The 6,976 DZs, 6 age ranges, and 2 sexes divide the population into 83,712 subpopulations, each with *∼*0–100 individuals and we term *cohorts*. We fit a Random Forest regression to cohort-level returning booster uptake. The model is informed by:

- age range;
- sex;
- ethnicity (% population belonging to a black or minority ethnicity), and;
- DZ-level deprivation ranks, by *access*, *income*, *employment*, *education*, *health*, *crime* and

*housing*.

To keep the model simple as well as suitably defined for generating scenarios with lower uptake (later in Section 4), we use DZ-level ranks rather than individual measures of deprivation as explanatory variables (these themselves have a strong degree of correlation, see Supplementary Material, Fig S1). Details of the individual deprivation ranks that feed into the model, chosen hyperparameters and performance are given in Supplementary Material, Section A.

The model explains 83.1% of between-cohort variation in returning uptake (fit: 85.6%, test: 74.2%), and 90.9% of variation between DZs (calculating uptake for all individuals aggregated by DZ) (fit: 92.7%, test: 83.6%) (Fig S2). There is evidence of clustering in residuals that mostly falls away past distance scales of order 5 km (as measured by the Moran’s I statistic [35] in Fig S3), suggesting variation in uptake from local factors beyond the variables used to inform the model.

Variable importance analysis indicates that all variables used serve to improve the model fit (Fig S4), with age the most important explanatory variable, consistent with the empirical trends (Fig S5). To associate a “directionality” to the effects of each of these variables we also conducted a partial dependence analysis (Fig S6), measuring the influence of different variable values on the average model outcome broken down by age group. Over the deprivation ranks, Education and Housing emerge as having the strongest effects, with a distinct drop-off with increasingly severe deprivation. This is more pronounced in younger cohorts for Education, but for older cohorts for Housing. The partial dependencies also indicate strongly that communities with higher black and minority ethnic (BAME) populations are on average associated with lower fit uptake. Per the census data, fewer than 11% of Scotland’s DZs have a BAME population over 25%. The data do not specify individual-level ethnicity so we can not infer a direct relation here, however the relation found is consistent with empirical studies finding lower relative uptake in BAME communities [36, 37, 38, 39]. Nonetheless, the inclusion of ethnicity (measured here as the percentage of a cohort belonging to a black or minority ethnicity) improves the model performance, with a high node purity (Fig S4) indicative of a stronger influence on cohorts where the 20+ BAME population significantly deviates from the DZ mean of 11%.

## 4 A method for predicting future distributions of uptake

Our Random Forest regression model reproduces detailed spatial patterns with high accuracy despite not being informed by spatial data explicitly (such as where DZs are located or nearest neighbours). The variation in returning first booster uptake from Fig 1A and as discussed in Section 3 indicate that we should expect any further falls in future rollouts to also be unequal across different demographics. We now explore whether the trends in our first booster model can provide clues as to which groups and geographic areas may be at highest risk of falls in uptake in the future.

Our regression model takes input data on deprivation and population structure. With our model we are free to feed in data that have been modified in some way, such as data where population structure is unchanged, but the profile of deprivation is different. In doing this, the model will produce uptake values that may differ from those using the “true” data. Such adjustments of the input data form the basis of standard machine learning methods such as feature importance where a variable is effectively “removed” under random shuffling, and partial dependencies (also *accumulated local effects*), where a single variable is modified in isolation to assess its influence on a model outcome [40].

Along these lines, we propose a novel method for assessing the risk of different population groups in suffering disproportionate falls in uptake in the future. Our hypothesis is that deprivation is the key driver for *spatial* differences in vaccine uptake (as evidenced in Figure 2), and that *population groups whose fitted values are more sensitive to small changes in community deprivation are prone to suffer disproportionately higher falls in uptake*.

We then adjust input data in a manner that *reduces* predicted booster uptake, and assess the resulting distribution. This expands on the partial dependence approach in two ways. First, instead of adjusting a single variable to assess the impact on the model outcome, we adjust all measures of deprivation (in this case, ranks) in parallel. Second, Random Forest models perform poorly when presented with values that exceed the data it was fit to (i.e. Random Forest models alone are poor at extrapolation), so we need a means of extending the model prediction for when a counterfactual deprivation rank falls below 1.

We detail our methodology in Supplementary Material, Section B. Referring to Fig 3, for each cohort we adjust the deprivation ranks of its associated DZ over a range of (both positive and negative) values of Δ, producing a set of predictions of uptake, and in turn determine how sensitive the predicted uptake is to changes in deprivation. The parameter Δ here is abstract, without a physical analogue. We then fit to each of these predictions a curve (a sigmoid function) as a function of Δ to these modelled values. By taking different values of Δ, we then extrapolate distributions of uptake under these counterfactual data. The effect of this functional description is to smoothly extrapolate the deprivation relationships to consider some nominal community with highly severe deprivation (i.e. that would rank below any existing DZ across all deprivation indices). As the value of Δ falls to larger negative values, the uptake over all cohorts will fall, but more sharply in cohorts that are more sensitive to changes in deprivation rank. Conversely, cohorts that are less sensitive to changes will have lower than average falls.

**Figure 3:**
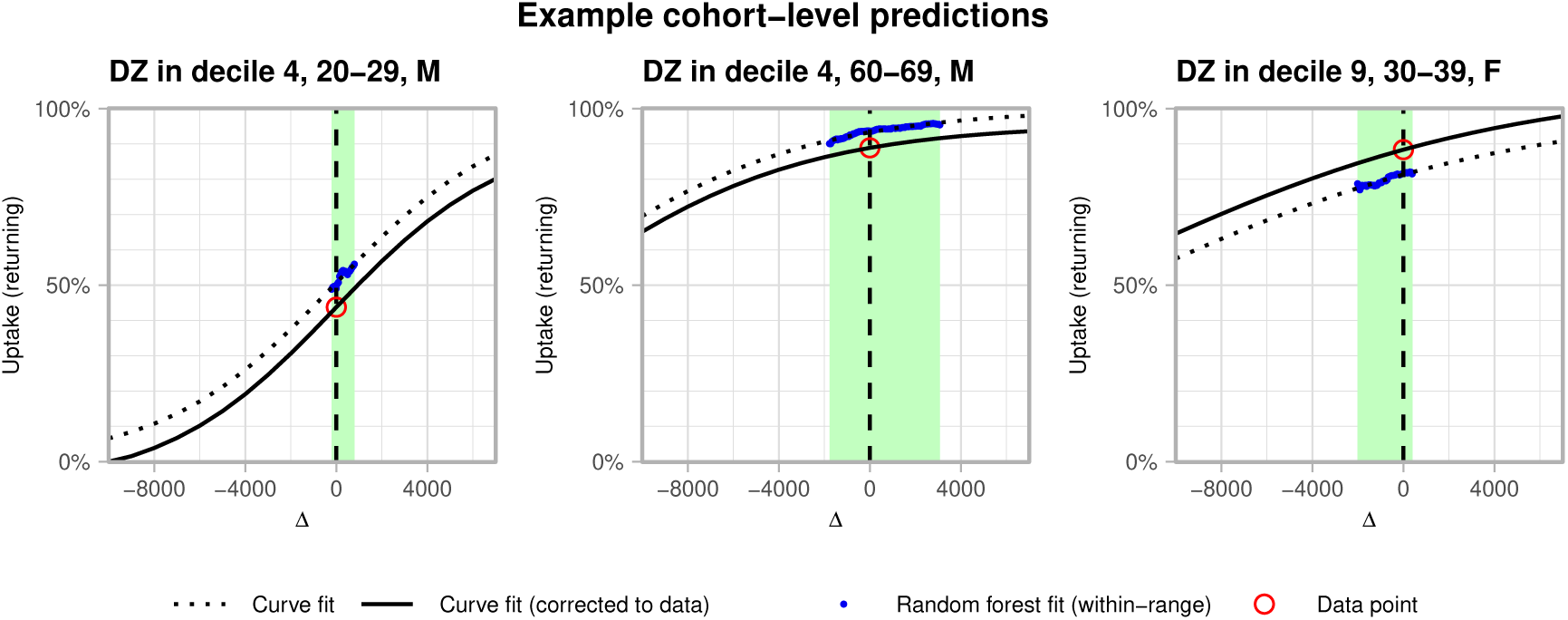
Graphical representation of prediction of uptake with respect to level of deprivation. For each cohort, the green box bounds the “floor” and “ceiling” values of deprivation rank shift Δ. Within this range, the projected uptake (blue points) falls for decreasing Δ (increasing level of deprivation). A sigmoid function (black, dotted) is fit to these fit values, which is then shifted to match the actual returning first booster uptake (red circle) at Δ = 0 (vertical dashed line).

### Prediction vs second booster in 75+, August 2022

To test the predictions of our model (fit to first booster uptake) in lower uptake scenarios, we first compare a model prediction to the distribution of second boosters in ages 75+, from a snapshot dated August 2022. Returning uptake for 75+ at this point was 82%. Fig 4 shows the skew with respect to deprivation is well captured, with a predicted ratio of uptake between the highest and lowest deprivation deciles of 1.16, compared to an actual ratio of 1.18.

**Figure 4:**
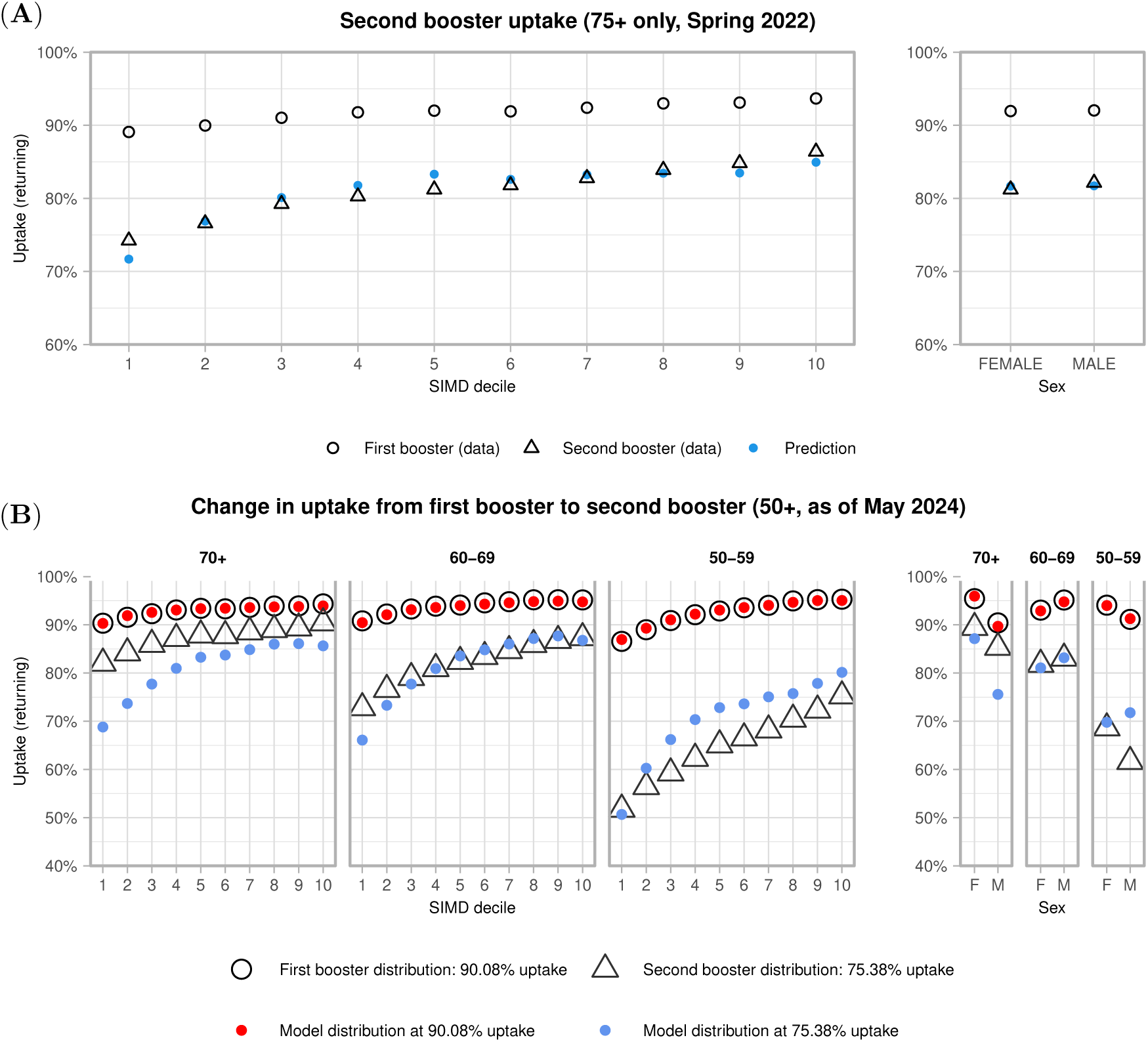
(**A**) Returning second booster uptake amongst individuals aged 75+, as of August 2022. Filled points are predictions from a model trained on first booster data, when matching the lower uptake in second boosters. (**B**) Returning uptake in the second booster rollout as of May 2024 across ages 50–59, 60–69, 70+, broken down by sex and deprivation decile, and the corresponding predicted model distribution when matching the second booster uptake of 75%.

### Prediction vs second booster in 50+, May 2024

With the current data, returning second booster uptake in ages 50+ is 75%, as compared to 90% for first boosters. An uptake of 75% corresponds to a value of Δ *≈ −*7290 (an 50+ uptake of 75% is predicted by our model if all DZs were *∼* 7290 ranks “lower”). Fig 4 shows our modelled distribution with respect to age, sex and deprivation as compared to actual uptake. The model predicts increased skews with respect to sex (ratio 1.04), age (1.16) and deprivation (1.39), relative to first booster uptake. However the predicted skew with respect to age is an underprediction (whereas in reality uptake fell especially sharply in 50–59), and overpredicts the skew with deprivation (whereas in reality the trend was less pronounced).

## 5 Discussion

We have used high-resolution data to describe patterns in COVID-19 booster vaccination uptake across communities in Scotland. First booster data reveal falls in uptake in younger, more deprived populations relative to the initial vaccination course. This is consistent with observed increased vaccine refusal in younger groups living in more deprived communities [41, 42, 36]. These inequalities in uptake then manifest as larger clusters of low uptake coinciding with communities with higher general deprivation. Data from the second round of booster vaccination show that as well as a general fall in uptake, skews seen in first boosters are exacerbated further, mainly by age. The distribution of second boosters represents uptake in the rollout furthest in time from the earlier acute phases of the pandemic.

To better understand fine-scale differences in first booster uptake, we have fit a Random Forest regression model, informed by local population structure and deprivation. This explains substantial spatial variation in returning booster uptake, and accurately captures differences by age, deprivation and sex. A community’s booster uptake can therefore be estimated with high accuracy solely from its population structure and relative level of deprivation, without information on where that community is physically located in Scotland, or who its neighbours are.

We then explored a method for predicting future distributions of uptake, using our first booster model. We use *prediction* here in the sense of prediction forecasting, i.e. to provide estimates of what might actually occur in the future, under the assumption that the underlying conditions of the prediction do not change [31]. We created uptake distributions using counterfactual data, where population structure was unchanged but each cohort had more severe deprivation than it does in reality, fitting a relation between estimated uptake and changes in deprivation. Population groups whose model prediction was more sensitive to small changes in deprivation are interpreted as being at increased risk of falls in uptake.

When set to predict the distribution for the single age group that was eligible for a second booster in Spring 2022, when conditions for vaccination were most similar to the first booster period (high awareness with an intensive campaign to boost uptake), the model successfully captures the increased skew with respect to deprivation. However when predicting the distribution of second boosters as of May 2024 across multiple age groups, the prediction is poor compared to the data, and over-estimates the skew with respect to deprivation, and under-estimates that with age, suggesting the underlying risk factors for vaccine refusal were *not* the same between first and second boosters. This is consistent with the changed context of the pandemic by the time second boosters were administered in the Autumn 2022; for first boosters, eligibility was rapidly expanded from ages 50+ to all adults in response to an outbreak of the B.1.1.529 *Omicron* variant of concern which saw the re-imposition of some non-pharmaceutical interventions especially in mass gatherings and hospitality [43, 44]. However throughout 2022 these were eased, including dropping the requirement for a vaccine passport in some hospitality settings [45], and global travel restrictions were gradually relaxed. The sharper drop-off with age than predicted by our model fit to first boosters is consistent with a broader shift in COVID-19 strategy towards protecting those deemed most vulnerable [46]. To probe the sensitivity of different cohorts to changes in deprivation using data within the range the model was trained on, we then proposed a method for extending to counterfactual data that effectively exceeds this range, which has provided useful insight. Such an extrapolation as implemented here is necessary as Random Forests perform very poorly with data beyond its training range. Nonetheless, this demonstrates how such a statistical model can be exploited in a novel way to explore counterfactual scenarios, and indicate when underlying risk factors may have changed.

The fall from first to second booster uptake combined with an increased inequality with deprivation highlights a potential *twofold* risk in the longer-term. A fall in uptake obviously reduces the amount of vaccine-induced protection against severe COVID-19 disease in the population. Compounding this though, with a growing skew in uptake, any shrinking pool of protection may become increasingly associated with those living in less deprived neighbourhoods, already at lower prior risk of developing severe disease. This more substantial fall in uptake from a lower baseline would see clusters of communities that are most vulnerable disproportionately more exposed, and at higher risk of infection spread, hospitalisation and mortality.

Finally, for the purposes of fine-scale infectious disease modelling, the profile of vaccine-induced immunity is one of few inputs that can be done accurately at the individual level with existing data. With early COVID-19 the exception, epidemiological data are typically sparser, and often representative of only a subset of infections that are severe (hospital/intensive care admissions, mortalities). We have shown here how such data can be used to inform how uptake may be modelled in the longer-term, under a set of assumptions. However, despite being routinely collected for many infectious diseases, these vaccination data are not often made available at such a fine spatial resolution to researchers. Our access in this instance has been exceptional, owing to the need for rapid, policy relevant analysis during the COVID-19 pandemic [47]. Our insights from these data otherwise has been entirely done using public data, using methods widely applicable to diseases beyond COVID-19, and to other indicators for vaccine hesitancy and accessibility beyond deprivation. With the wealth of public demographic data available, our results provide a strong argument for the value of access to such vaccine uptake data for COVID-19 as well as other infectious diseases where there is a strong public health interest.

## 6 Author contributions

R.R.K. conceived the project. A.J.W. wrote the model code, performed the model analysis, and with R.R.K. wrote the manuscript. A.M.M. and M.S. motivated conceptual work on evolving motives for vaccination, and risk factors for declining uptake. R.R.K. and A.J.W. conceived the technique for generating lower-uptake scenarios. All authors commented on and approved the manuscript.

## 8 Code availability

Model code is available at https://git.ecdf.ed.ac.uk/awood310/covid-19-vaccination-analysis-and-project

## 9 Data availability

The vaccination data utilised in this work are not publicly available. They are provided to the authors for academic research by Public Health Scotland’s electronic Data Research and Innovation Service, under a data sharing agreement (“Spatial and Network Analysis of SARS-Cov-2 Sequences to Inform COVID-19 Control in Scotland”).

Two versions of the vaccination data have been used in this analysis; the first specifying at a pseudonymised individual level, each individual vaccination event up to 18 August 2022, which produced Figure 4. The authors no longer have access to this version after expiry of the DSA. The second version used for all other analyses as detailed in Section 2 is a version aggregated into age/sex/DZ population groups, giving time-aggregated doses as of May 2024. As of September 2024 this DSA remains active. All other data utilised in this work are publicly available drawing primarily from Scottish census data.

## Supplementary Material

### A Data and model details

The eDRIS data specify for each DZ/age range/sex population group, the total number of individuals to have received exactly one dose, two doses, three doses and four doses. When that is fewer than five individuals, the exact number is not given. To estimate a number of individuals when it is fewer than five, for each population group we fit a two-parameter gamma distribution to the number of individuals to have received exactly *n* doses, extrapolating to the *<*5 range. We then estimate the number of individuals for each *<*5 entry by drawing a value from the distribution.

The random forest model was fit using the *RandomForest* package (version 4.6–14) in *R* (version 4.1.0). Model code is available at https://git.ecdf.ed.ac.uk/awood310/covid-19-vaccination-analysis-and-projection The ranking of DZs by deprivation are taken from the Scottish Index of Multiple Deprivation, which is publicly available at https://simd.scot alongside a detailed methodology of how DZs are ranked. Briefly here, DZs are ranked by deprivation across each of:

- *Access*: an aggregate of measures including access to high-speed broadband, and the average time it takes to travel to public services either by car or public transport;
- *Crime*: the rate of recorded crimes per population;
- *Employment* : relating to the proportion of working adults who are either not working, or in receipt of a type of employment benefit;
- *Housing* : based on the proportion of individuals living in households that are classed as overcrowded, or without central heating;
- *Education*: an aggregate of measures including as the proportion of working age people with no qualifications, and school pupil attendance;
- *Income*: based on the proportion of individuals that are considered *income deprived*, such as being in receipt of Universal Credit;
- *Health*: an aggregate of measures of poor health outcomes including hospital stays due to drug or alcohol misuse, and the number of emergency stays in hospital relative Scotland as a whole.

We trained a set of models with different hyperparameters, testing across: number of variables tested per tree split (2, 3, 4, 5), maximum node size (2000, 3000, 4000), and random number seed (7 seeds tested each), and training proportion (70%, 80%). Each model had 1000 trees. We chose the hyperparameters and seed of the model (node size 5000, training proportion 80%, max node size 4000) that explained the most DZ-level variation (R-squared) in the DZs the model was not trained on.

**Figure S1:**
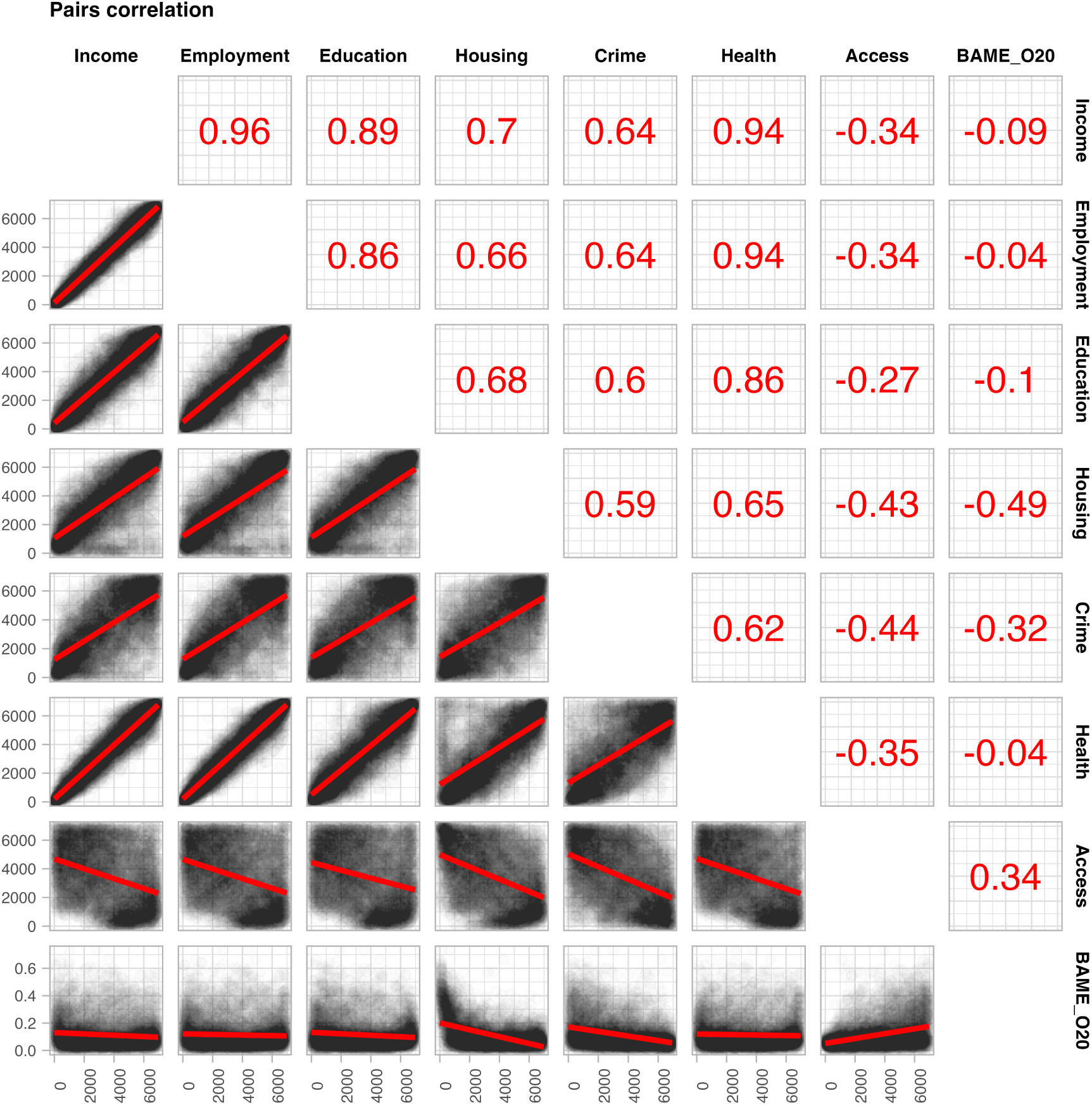
Correlation plot (at DZ level) between the different deprivation ranks, as well as the proportion of residents aged over 20 belonging to a black or minority ethnicity.

**Figure S2:**
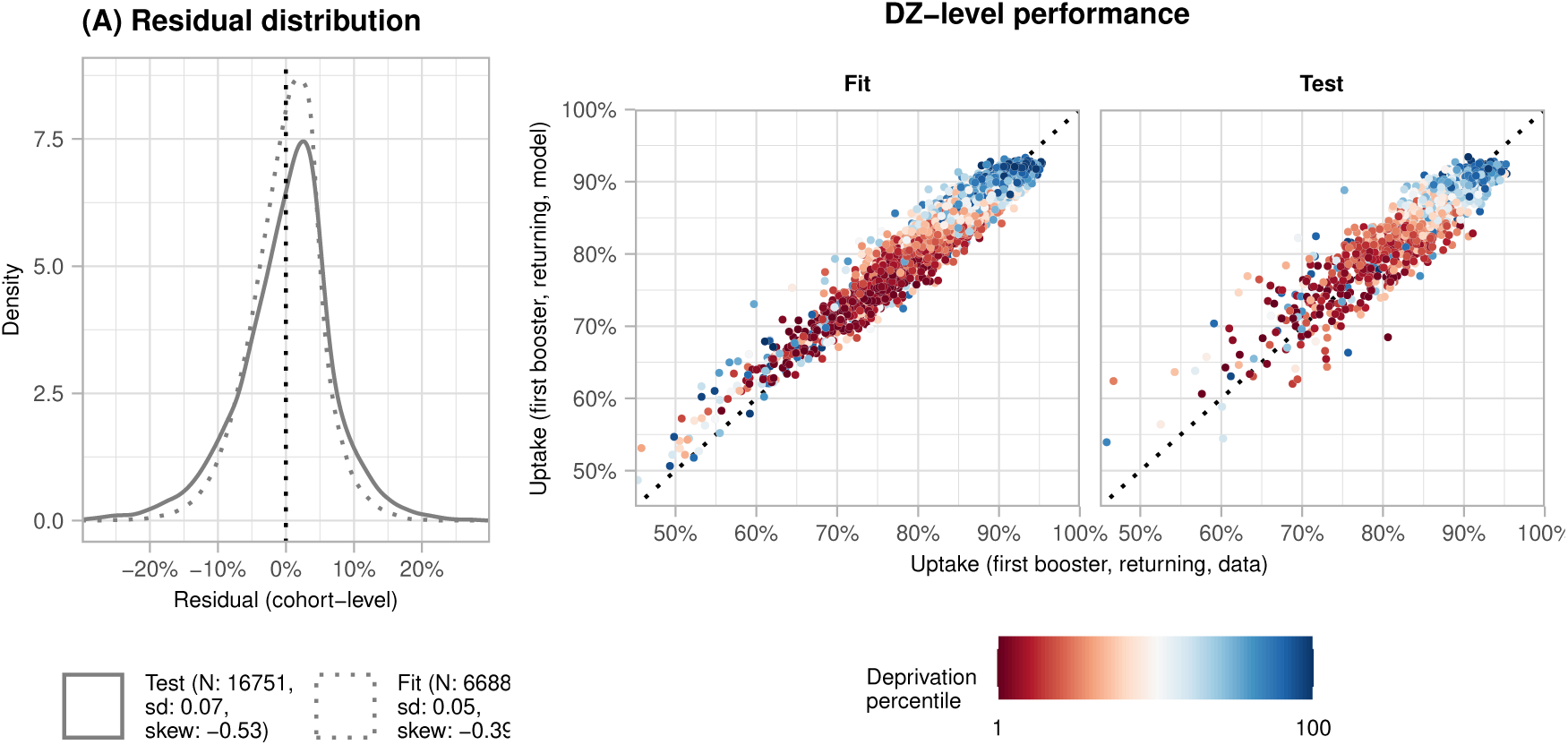
Model performance. (**A**) Residual distributions of the fit and test data sets. (**B**) Performance comparing data and fit values for returning uptake at the DZ level, over individual DZs, with deprivation indicated.

**Figure S3:**
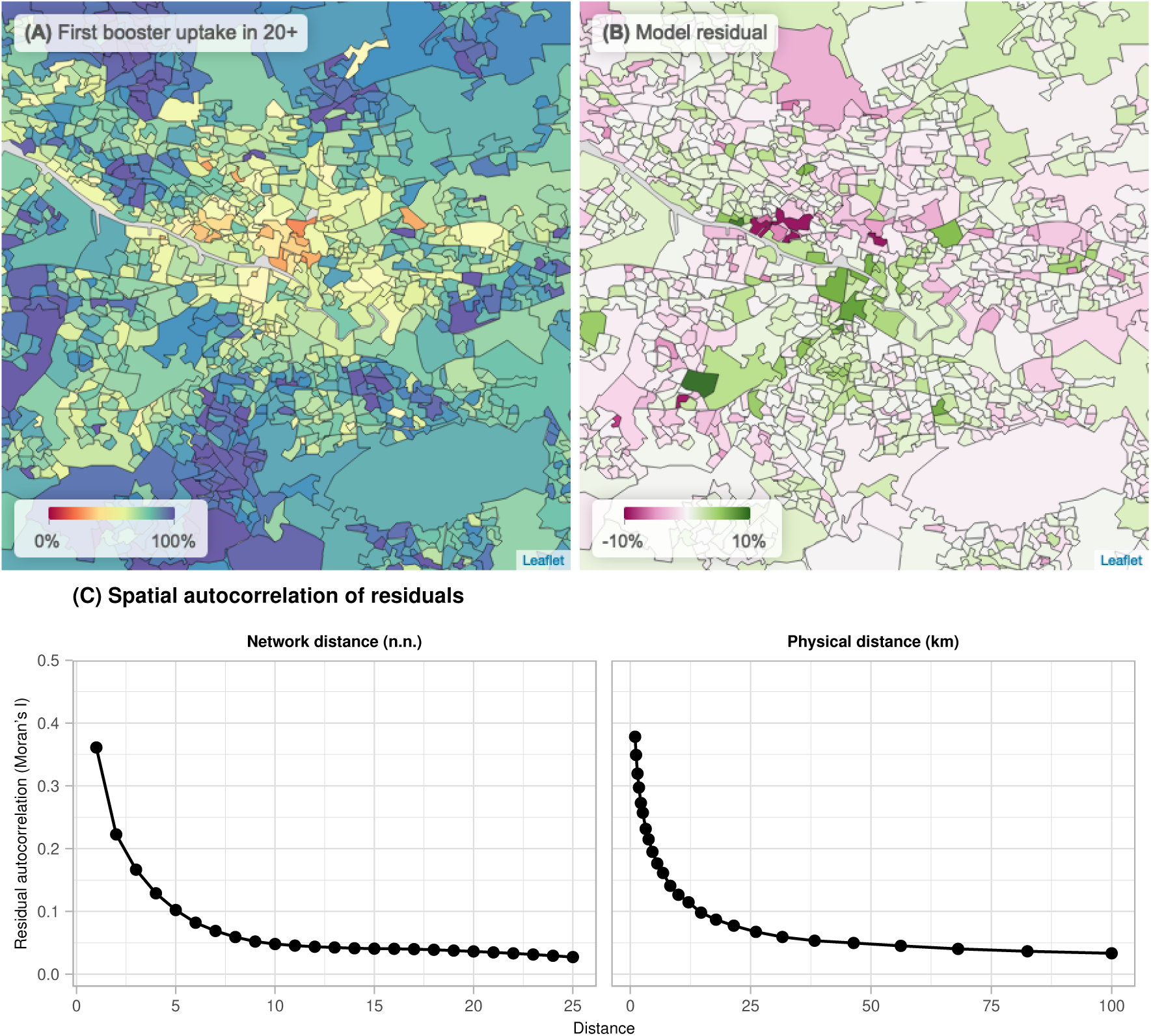
(**A**) overall booster uptake across an 18km *×* 18km area of central Glasgow, and (**B**) corresponding residuals (the difference between actual uptake, and the fit value). Green DZs are those where observed uptake is higher than the fit value, and pink DZs indicate where observed uptake was lower. (**C**) Residual autocorrelation as measured by the Moran’s I statistic, comparing autocorrelation between residuals (*y*-axis) within a certain locus (*x*-axis). Autocorrelations fall substantially past distances of 5–10km, or within 1–5 nearest neighbours (with nearest neighbours being DZs that share a border).

**Figure S4:**
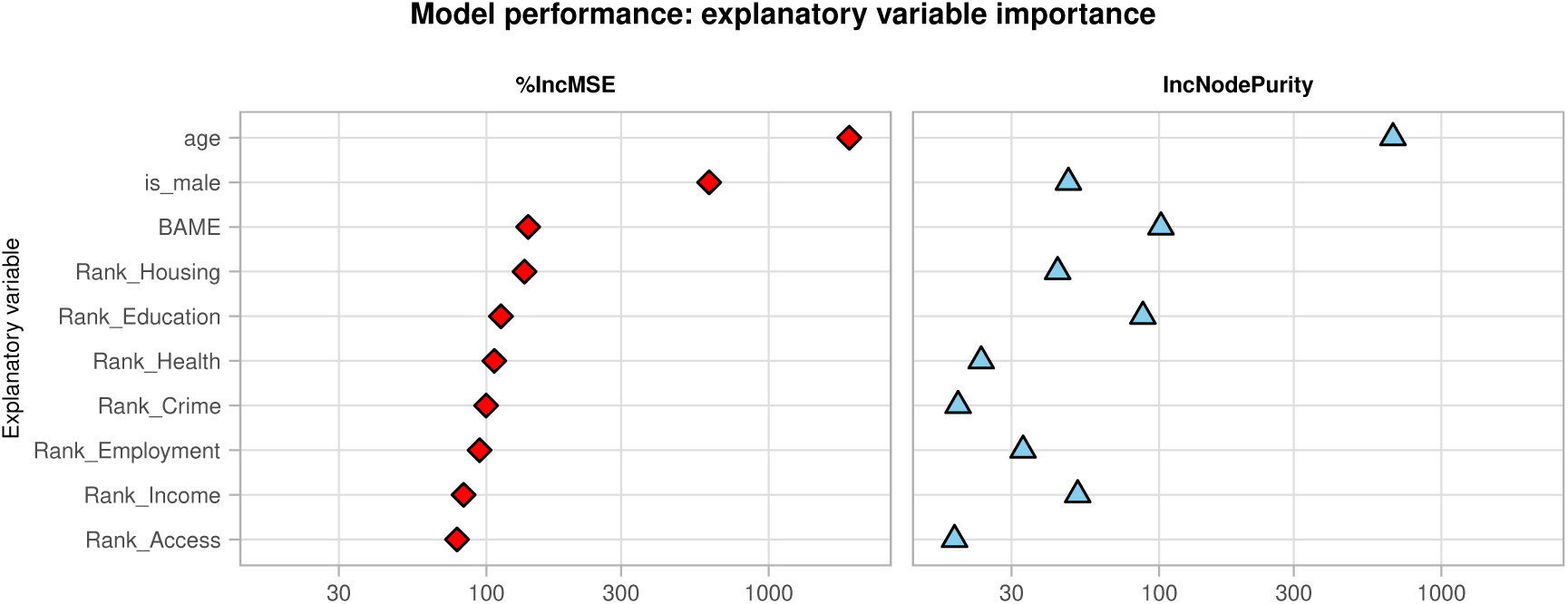
Explanatory variable importance output from RF model (MSE loss and node purity).

**Figure S5:**
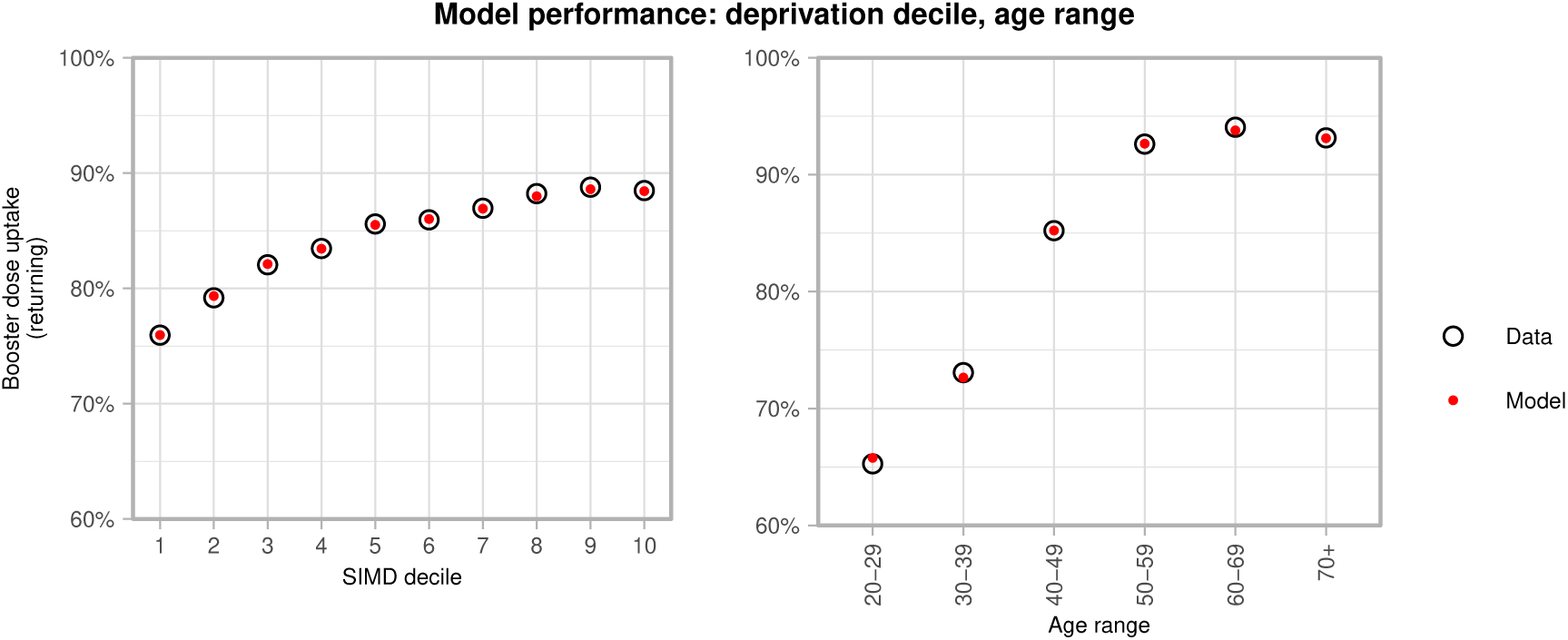
Model performance aggregating cohorts over deprivation decile, and age range.

**Figure S6:**
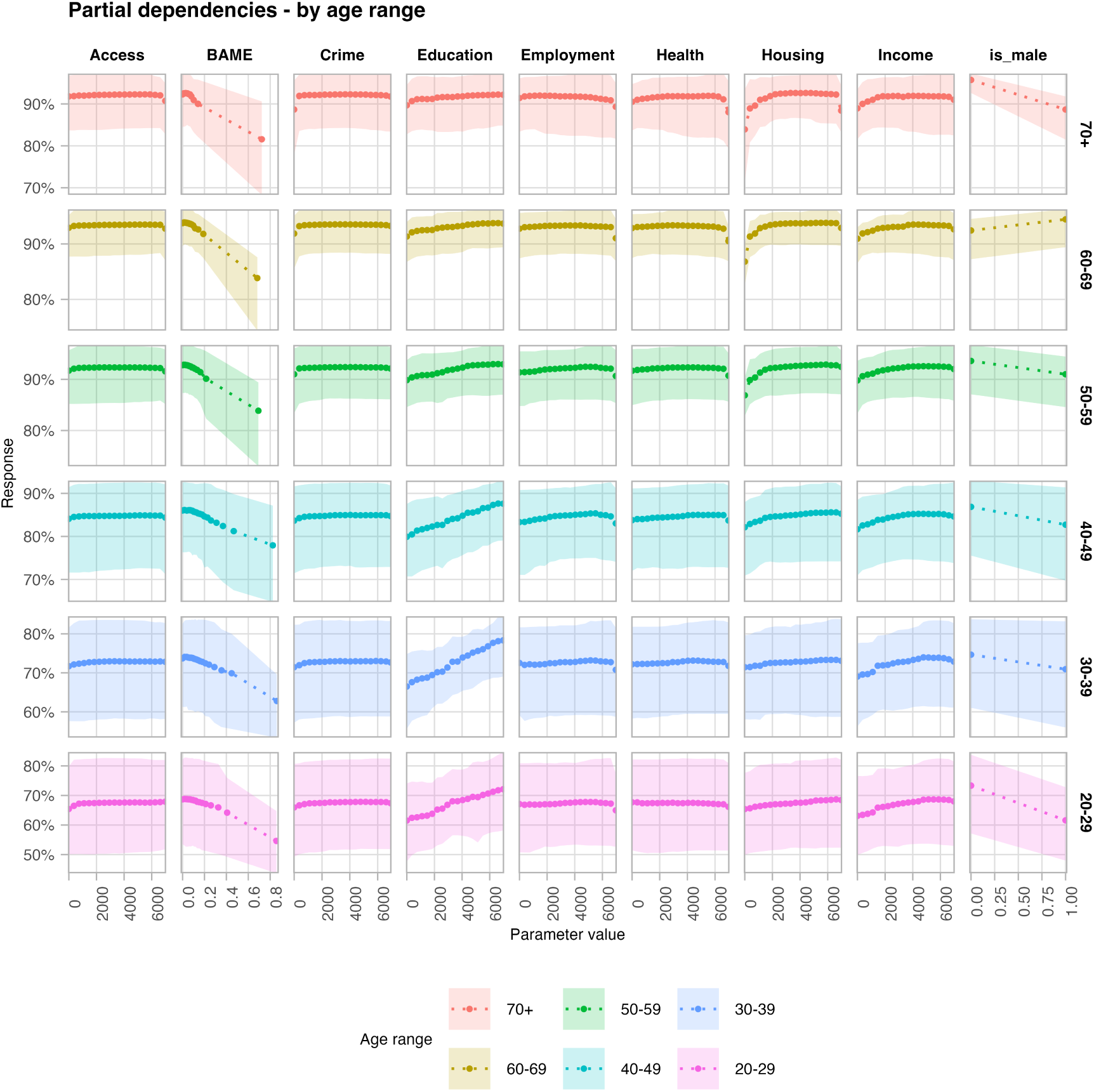
Partial dependence plots from the random forest regression divided by age range. This indicates the average “response” (the model prediction) to each of the particular parameter values, averaged over many different cohorts. The filled region indicates the central 90% of responses.

### B Details of modelling lower-uptake uptake distributions

We use uptake in the first booster vaccination rollout, in conjunction with our random forest model informed with SIMD deprivation ranks, to create estimates of longer term distributions, with *lower* nationwide uptake.

Per Section 4, we create counterfactual *input* data by making a univariate shift of all deprivation ranks in dataset by some amount Δ. For larger negative values of Δ (thus the ranks being lower, and representative of higher levels of deprivation than in reality), projected uptake is anticipated to fall. A limitation to this is when a counterfactual rank runs outside the range 1–6,976 (e.g., a rank 55 at shift Δ = *−*500 will be “rank *−*445”). Random forest models, being a form of stepwise regression, generally perform poorly with data outside the range they are fit against. To address this and project beyond the “floor” rank of each DZ, then, we fit a sigmoid function to each cohort *i*, for the fit returning uptake *U_i_* as a function of Δ

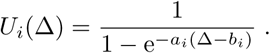

*a_i_* and *b_i_* are parameters to be fit for each cohort, based on the model fit values for the range of counterfactual Δ values that do not exceed the range 1–6,976. Finally, we shift each fit curve along the *y*-axis by an amount *ɛ_i_*, such that *U_i_*(0) is the exact observed first booster uptake for cohort *i*. To avoid scenarios where this shift introduces a modelled uptake greater than 100% or below 0%, we bound 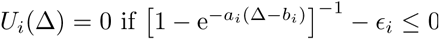, and 1 if 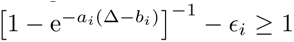. The Δ = 0 distribution then exactly reproduces that observed for first boosters. We then reduce Δ to generate predicted, lower-uptake distributions.

For about 13% of cohorts the range of valid values for Δ is too narrow to reasonably fit a curve (i.e., instances where a DZ has one very high deprivation rank, and another that is very low). For cohorts where the range is lower than 500 ranks, then, we instead use the average value of parameters *a_i_* and *b_i_* from similar cohorts; having the same age range, sex, DZ deprivation *decile*, and similar first booster uptake.

The underlying random forest model is fit to returning uptake rather than overall uptake. We then calculate the corresponding overall uptake by multiplying relative uptake by overall first dose uptake. By making lower-uptake distributions by decreasing returning uptake, then, we make an implicit assumption that all future doses will be given to individuals that have received at least one vaccination before.

